# Mode and dynamics of *vanA*-type vancomycin-resistance dissemination in Dutch hospitals

**DOI:** 10.1101/2020.07.21.20158808

**Authors:** Sergio Arredondo-Alonso, Janetta Top, Jukka Corander, Rob J L Willems, Anita C Schürch

## Abstract

**Background:** *Enterococcus faecium* is a commensal of the gastrointestinal tract of animals and humans but also a causative agent of hospital-acquired infections. Resistance against glycopeptides and especially to vancomycin, a first-line antibiotic to treat infections with multidrug-resistant Gram-positive pathogens, has motivated the inclusion of *E. faecium* in the WHO global priority list. Vancomycin resistance can be conferred by the *vanA* gene cluster on the transposon Tn*1546*, which is frequently present in plasmids. The *vanA* gene cluster can be disseminated clonally but also horizontally either by plasmid dissemination or Tn*1546* transposition between different genomic locations. Here, we reconstructed all nested genetic elements (clone, plasmid, transposon) to study how the dissemination of *vanA*-type vancomycin resistance occurred in Dutch hospitals (2012-2015).

**Methods:** We performed a retrospective study of the genomic epidemiology of 309 vancomycin-resistant *E. faecium* (VRE) isolates across 32 Dutch hospitals (2012-2015). Genomic information regarding clonality and Tn*1546* characterisation was extracted using hierBAPS sequence clusters (SC) and TETyper, respectively. Plasmids were predicted using gplas in combination with a network approach based on shared k-mer content. This allowed determining all nested genomic elements (clone, plasmid and transposon) involved in the dissemination of the *vanA* gene cluster. Next, we conducted an “all vs. all” pairwise comparison between isolates sharinga potential epidemiological link to elucidate whether clonal, plasmid or Tn*1546* spread accounted for the dissemination of *vanA* resistance.

**Results:** The 309 VRE isolates belonged to 18 different SCs of which SC13 (n = 102, 33%), SC17 (n = 52, 16.8%) and SC18 (n = 42, 13.6%) were predominant. We identified seven different plasmid types bearing the *vanA* gene cluster, four of which were highly similar (identity ∼99%, coverage∼84%) to previously described complete plasmid sequences. We estimated that clonal dissemination contributed most (∼45%) to the spread of vancomycin-resistance in Dutch hospitals, followed by Tn*1546* mobilisation (∼12%) and plasmid dissemination (∼6%).

**Conclusions:** The dissemination of the *vanA* gene cluster in Dutch hospitals between 2012 and 2015 was dominated by clonal spread. However, we also identified outbreak settings with high frequencies of Tn*1546* transposition and/or plasmid dissemination in which the spread of resistance was mainly driven by horizontal gene transfer (HGT). This study demonstrates the feasibility of distinguishing between modes of dissemination with short-read data and provides one of the first quantitative assessments to estimate the relative contribution of nested genomic elements in the dissemination of *vanA-*type vancomycin resistance cluster.

## Background

*Enterococcus faecium* is commonly inhabiting the gut of animals and humans but has also emerged as a nosocomial pathogen causing a sizable fraction of healthcare-associated infections, specifically device-associated infections like central line-associated bloodstream and surgical site infections (1,2). The intrinsic and acquired multi-drug resistance against fluoroquinolones, aminoglycosides and more importantly against glycopeptides motivated the inclusion of *E. faecium* in the WHO global priority list (3). The number of strains resistant against vancomycin, a first-line glycopeptide antibiotic to treat infections with multidrug resistant Gram-positive pathogens, dramatically increased first in the US in the 1990s, followed by other parts of the world (4). Resistance against vancomycin can be acquired through eight different gene clusters (*vanA, vanB, vanD, vanE, vanG, vanL, vanM*, and *vanN*) (5,6) of which *vanA* and *vanB*, associated to transposon sequences Tn*1546* and Tn*1549*, respectively, are the predominant vancomycin-resistance gene clusters (7).

Clonal spread of vancomycin-resistant *Enterococcus faecium* (VRE) has been extensively described using a plethora of molecular typing schemes. They range from fingerprint-based methods like pulsed-field gel electrophoresis (8), to PCR-based methods such as multiple-locus variable number tandem repeat analysis (9), multilocus sequence typing (10) and whole genome sequencing (11). However, due to the fact that vancomycin resistance genes are located on mobile genetic elements, vancomycin resistance also has the potential to be transferred horizontally. In fact, mobilization of the *vanA* gene cluster via insertion in different plasmid backbones has already been reported (12,13). To identify the dissemination of *vanA* plasmids within hospital settings, whole-genome sequencing (WGS) based on short-read technologies has been recently applied to collections of hundred hospitalized patient isolates in Denmark and Australia (14,15). These studies undertook a reference-based approach to map short-reads against complete plasmids from a selection of isolates. However, this approach can overestimate the presence of a reference plasmid by neglecting the mosaicism observed in these types of sequences as previously observed for *Enterobacteriaceae* isolates (16) and *Enterococcus* populations (17).

The dissemination of the *vanA* gene cluster can occur vertically, in which case the same plasmid type and Tn*1546* variant are observed in two clonal isolates. However, the *vanA* gene cluster can also be horizontally transferred, by two different processes: i) plasmid dissemination which is reflected by observing the same plasmid type and Tn*1546* variant in strains that have a different clonal background, and ii) transposition of Tn*1546* between different plasmid types (18–20). This nested nature of these mobile genomic elements resembles the Russian-Doll model which has been previously used to describe the transfer of carbapanamese genes, *bla*^kpc^, in *Enterobacteriaceae* (*16*). *It is important to note that the Tn1546* is a non-conjugative transposon but its mobilisation can occur when the element is embedded in another conjugative element.Furthermore, the presence of IS elements (e.g. IS*1216*) surrounding the transposon can mobilise the *vanA* gene cluster to other genomic locations (18–20).

The genomic approach conducted here allowed to fully reconstruct and quantify the most likely mode of dissemination by characterizing the clonal background (hierBAPS SC), *vanA* plasmid type (*de novo* prediction and network assignment) and T*n1546* variants harboring the *vanA* resistance gene cluster. Clonal dissemination, defined by vertical inheritance of the same SC, *vanA* plasmid type and Tn*1546* variant, was the most frequent scenario of vancomycin-resistance dissemination occurring in the Netherlands between 2012-2015. However, we also detected outbreak settings in which Tn*1546* transposition between distinct plasmid types and/or plasmid dissemination were the dominant mechanisms driving the *vanA* gene cluster dissemination. To our knowledge, we provide one of the first studies to estimate the frequencies of clonal and HGT processes in the dissemination of the *vanA* gene cluster occurring in Dutch healthcare settings between 2012-2015.

## Methods

### Dutch VRE collection, short-read WGS and genome assembly

The isolates from this collection represent a subset of isolates from a previous study we conducted and that consisted of 1,644 *E. faecium* isolates (21). Isolates with the v*anA*-type vancomycin-resistance gene cluster (n=309) from 32 Dutch hospitals collected between 2012 and 2015 were further analyzed. DNA extraction, and whole-genome sequencing using Illumina NextSeq were conducted as previously described (22). Short-reads were trimmed using Trim Galore (version 0.6.4_dev) using the flag ‘--paired’ and specifying a phred score of 20 with the flag ‘--quality’ (23). We used Unicycler (version 0.4.7) passing the short paired-end trimmed reads from Trim Galore and specifying the normal mode (--mode) (24). Unicycler was used to compute the assembly graph provided in the file ‘assembly.gfa’ which selects for the k-mer size that optimises the ratio between number of dead-ends and contig size in the graph given by SPAdes (version 3.14.0) (25). In-silico prediction using Abricate (https://github.com/tseemann/abricate, version 0.8), with the ResFinder database (indexed on 16th of July 2018) (26) was conducted to search and select for isolates bearing the *vanA* resistance gene.

### Population structure of VREfm isolates

Recombination events and estimation of sequence clusters using BratNextGen and hierBAPS were performed as previously described (21). PopPUNK (version 2.0.1) was run specifying the flag ‘--easy-run’ with a minimum k-mer size of 13 (flag --min-k) and creating the files required to generate a microreact project (flag --microreact) (27).

### *De-novo* plasmid prediction

Gplas (version 0.6.1) was used to *de novo* predict the plasmids present in the assembly graph of each VRE isolate (28). Gplas was run using mlplasmids (22) as classifier to predict plasmid sequences (flag ‘-c’), specifying the species model ‘Enterococcus faecium’ (flag -s), a modularity threshold of 0.1 to partition the resulting bins (flag -q), and 50 walks per plasmid seed (flag -x).

### Tn*1546* characterisation

TETtyper (unique version) was used (29) to detect SNPs and deletions present in the Tn*1546* sequences of each complete plasmid sequence or predicted *vanA* plasmid bin against a reference sequence (--ref) corresponding to the original transposon structure (NCBI Nucleotide Accession M97297) described by Arthur et al. (30). We passed the trimmed reads to TETyper with otherwise default parameters.

### Network of complete plasmid sequences and plasmid type definition

Mash (version 2.2.2) (31) specifying a k-mer size (-k) of 21 and sketch size (-s) of 1,000 was used to perform k-mer pairwise comparisons between the 26 complete vancomycin-resistant (*vanA*) plasmids from the same collection of 1,644 *E. faecium* isolates (21). Based on the density distribution of Mash distances, we estimated an optimal cutoff of 0.025 to define the minimum distance to draw an edge between two nodes (complete plasmid sequences) in a network. The igraph R package (version 1.2.4) was considered to represent the network (32). Independent components (subgraphs, size > 1 node) in the network were considered as plasmid types (A-F).

To perform average nucleotide identity (ANI) measures between the complete plasmid sequences, we used the script ‘average_nucleotide_identity.py’ provided in the pyani tool (version 0.2.10) (33) with default parameters. We further considered the reported ANIm alignment coverage and ANIm identity values to support plasmid type assignments. The hierarchical clustering given by pyani, based on ANIm alignment coverage, was compared against the plasmid types defined using our proposed network approach.

For visualization purposes, the starting coordinate position of complete *vanA* plasmids was adjusted using the function fixstart from circlator (version 1.5.5) using a customized database of known plasmid replication initiator sequences (34). Easyfig (version 2.2.2) (35) with a minimum 80% identity and minimum block length of 500 bp were considered to visualize the blastn alignment (36) produced using the complete plasmid sequences belonging to the plasmid type B.

### Network of predicted plasmid bins and integration of plasmid types

The predicted plasmid bins reported by gplas and bearing the *vanA* gene cluster were pairwise compared using Mash (k = 21, s = 1,000). The igraph R package (version 1.2.4) was used to represent a network in which nodes corresponded to *vanA* plasmid bins and edges to connections between bins with a Mash distance lower than 0.025. The same threshold (0.025) to define an edge was considered since the density distribution of Mash distances followed the same pattern as previously observed with the complete plasmid sequences. The network consisted of 270 nodes and 16 independent components (subgraphs with > 1 node). Component 3 (236 nodes) was partitioned into 3 different graph groups based on its modularity value (0.42) using the function ‘cluster_louvain’ from the igraph R package (version 1.2.4). We focused on components/graph bins with > 10 isolates that were termed as plasmid bin groups (1-8).

Next, we integrated the plasmid types (A-F) into the network of predicted plasmid bins. For this purpose, we computed Mash distances (k = 21, s = 1,000) between complete plasmid sequences and predicted plasmid bins. The igraph R package (version 1.2.4) was used to represent a network in which nodes either corresponded to *vanA* plasmid bins or complete plasmid sequences and edges to connections between sequences (bins or complete sequences) with a Mash distance lower than 0.025. Plasmid bin groups with edges connecting to complete plasmid sequences were assumed to carry the same plasmid type (A, B, C, D). Plasmid bin groups without edges connecting to complete plasmid sequences were considered as carrying novel plasmid types (G, H, I).

### Contribution of nested genomic elements in the dissemination of *vanA*-type gene cluster

Pairwise comparisons were computed between VRE isolates sampled within 12 consecutive months and isolated at: i) same hospital, ii) same Dutch region and iii) country-wide. We determined which genomic elements were shared between pairs of VRE isolates and defined the following scenarios: i) clonal dissemination, characterized by identical SC, *vanA* plasmid type and Tn*1546* structure; ii) HGT plasmid dissemination, characterized by identical *vanA* plasmid type and Tn*1546* structure but distinct SC type, iii) HGT transposon dissemination,characterized by identical Tn*1546* structure but distinct SC and *vanA* plasmid types and iv) no linkage (unrelated cases), distinct Tn*1546* structure.

### Visualization of genomic elements

The R package maps (version 3.3.0) in combination with the R package ggplot2 (version 3.1.0) were considered to plot spatial information of the isolates specifying the map of the Netherlands. The R package genoplotR (version 0.8.9)(37) was used to visualize the gene structure and to highlight the *vanA* gene cluster location and the presence of IS elements present in the plasmid types (A-F). The R package ggtree (version 1.14.6) (38) was used to integrate the neighbour-joining tree based on the core-genome given by PopPUNK together with SC assignment and predicted *vanA* plasmid types. In addition, a Microreact project (version 15.0.0) (39) was created to integrate and visualize the genomic and metadata information.

## Results

### The population structure of VRE from Dutch hospitals

This study was conducted with samples from an extensive collection of 1,644 *E. faecium* isolates derived from clinical and non-clinical sources with associated short-read WGS data (21). We focused on Dutch clinical isolates with complete metadata information regarding clinical settings and isolation date, from 2012 to 2015 (n = 593). From this selection, 309 (52.1%) and 265 (44.7%) isolates carried the *vanA* and/or *vanB* gene clusters, respectively. We focused on the *vanA* VRE samples since the resistance gene cluster is frequently present on plasmids (40,41). This permitted us to investigate a nested genomic system in which the glycopeptide resistance can be disseminated on a clonal, plasmid and/or transposon level.

The clonality of these 309 *vanA* VRE samples was determined using hierBAPS (42), after filtering for recombination events as previously described (21). HierBAPS defined 18 different sequence clusters (SCs) of which SC13 (n = 102, 33%), SC17 (n = 52, 16.8%) and SC18 (n = 42, 13.6%) represented the most predominant clones in the dataset (Figure 1A). The distribution of these SC across time and geographical position showed that SC13 was widespread in Dutch hospitals for the entire collection period (2012-2015) (Figure 1B) compared to SC17 which was observed in distinct regions (Amsterdam, Lelystad, Zwolle) from August-September 2012 (Figure 1B). SC18 was detected around 2014 in several Dutch regions (Figure 1A). A core-genome based neighbour-joining tree of the samples was computed using PopPUNK (27), and it was integrated with metadata information in the following Microreact project https://microreact.org/project/FCUD_d1zt. Metadata information and SC assignment of the isolates are also available in Additional File S1.

**Figure 1.**
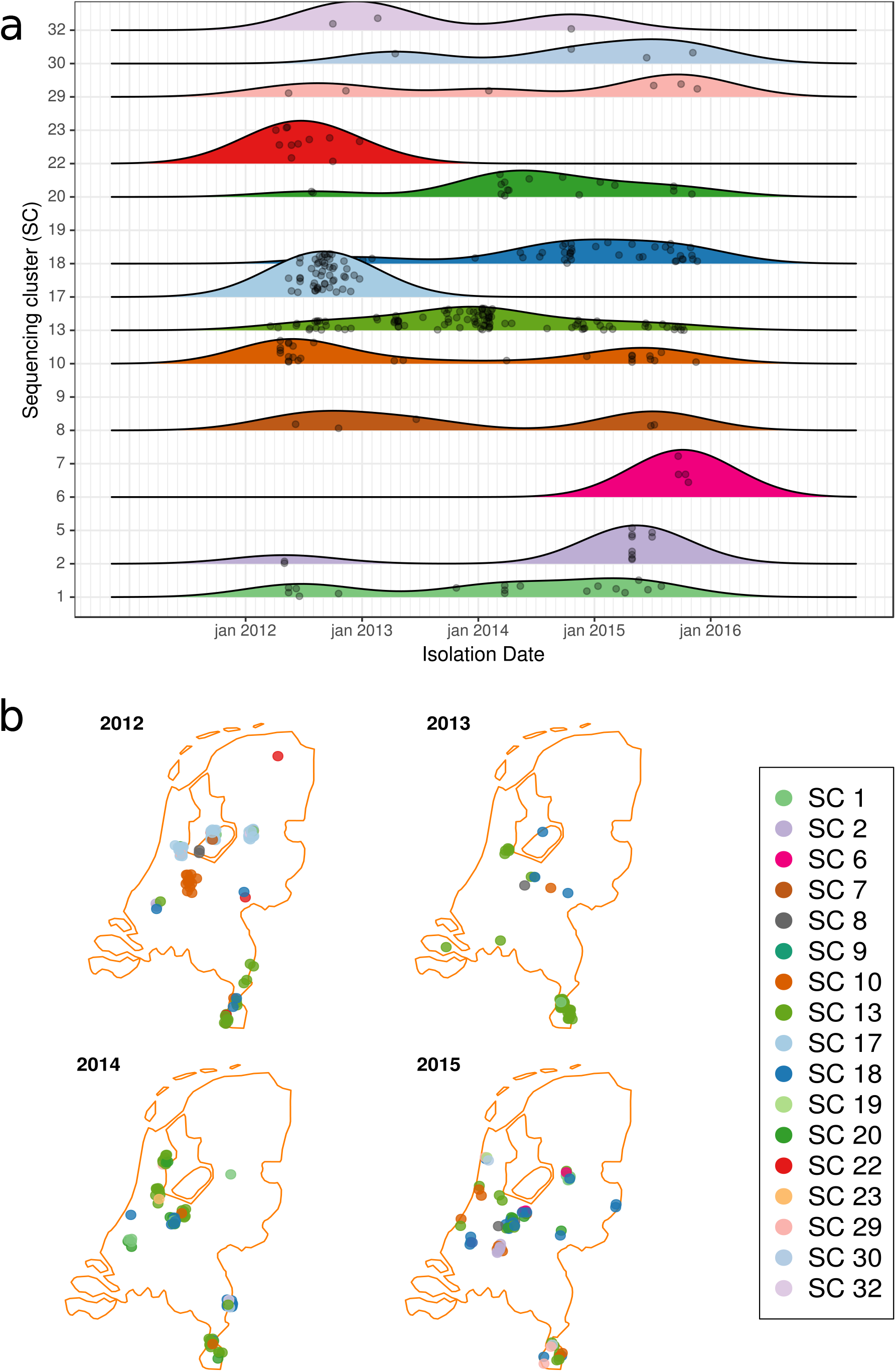
Temporal and spatial distribution of the 309 VRE isolates. A) Isolation date distribution of the sequencing clusters (SC) found in the 309 VRE isolates. B) Geographical distribution of the 309 VRE isolates along the years (2012-2015).

### Developing a novel plasmid typing scheme based on network clustering

To establish a partitioning scheme similar to hierBAPS SCs but uniquely based on the similarity between plasmids carrying the *vanA*-type gene cluster, we first retrieved 26 *E. faecium* complete *vanA* plasmids from the same collection of 1644 clinical and non-clinical isolates (21) with known isolation date and country (Table 1). These complete sequences were pairwise compared using Mash (k = 21, s = 1,000) and integrated into a network. Based on the k-mer distance distribution (Fig. S1), we defined an optimal cutoff of 0.025 to define an edge in the network. This resulted in six independent subgraphs in the network that signified distinct *vanA* plasmid types (A-F) with similar content and structure (Fig. 2A). Two complete plasmid sequences (E8172_3 and E8202_3) remained as singletons in the network and were not further considered in the analysis.

**Table 1.** Metadata information of the *vanA* complete plasmid sequences (n = 24) and average pairwise alignment coverage between and within plasmid types.

| Plasmid ID | Plasmid type | Country | Isolation Year | Isolation source | SC type | Between coverage <sup>a</sup> | Within coverage <sup>b</sup> |
| --- | --- | --- | --- | --- | --- | --- | --- |
| E0139_2 | A | the Netherlands | 1996 | Non-hospital | 29 | 40% | 85% |
| E0595_2 | A | the Netherlands | 1996 | Pig | 30 |  |  |
| E0656_2 | A | the Netherlands | 1999 | Non-hospital | 30 |  |  |
| E6975_4 | B | Greece | 2010 | Hospital | 13 | 32% | 85% |
| E7114_4 | B | Latvia | 2010 | Hospital | 13 |  |  |
| E7160_5 | B | Slovenia | 2009 | Hospital | 5 |  |  |
| E7240_4 | B | Greece | 2010 | Hospital | 10 |  |  |
| E7246_6 | B | Greece | 2009 | Hospital | 13 |  |  |
| E7471_5 | B | the Netherlands | 2012 | Hospital | 22 |  |  |
| E8423_3 | B | the Netherlands | 2015 | Hospital | 18 |  |  |
| E7313_3 | C | the Netherlands | 2012 | Hospital | 2 | 38% | 80% |
| E8014_3 | C | the Netherlands | 2014 | Hospital | 13 |  |  |
| E8414_4 | C | the Netherlands | 2014 | Hospital | 1 |  |  |
| E6055_4 | D | Portugal | 2010 | Hospital | 22 | 35% | 75% |
| E8040_4 | D | the Netherlands | 2014 | Hospital | 1 |  |  |
| E4227_3 | E | Sweden | 2005 | Chicken | 35 | 28% | 96% |
| E4239_3 | E | Sweden | 2007 | Chicken | 35 |  |  |
| E6020_3 | F | Latvia | 2010 | Hosp. | 8 | 38% | 82% |
| E6988_5 | F | Latvia | 2010 | Hosp. | 21 |  |  |
| E7025_5 | F | Latvia | 2010 | Hosp. | 23 |  |  |
| E7040_5 | F | Latvia | 2010 | Hosp. | 21 |  |  |
| E7067_5 | F | Latvia | 2010 | Hosp. | 21 |  |  |
| E7070_9 | F | Latvia | 2010 | Hosp. | 21 |  |  |
| E7207_6 | F | Greece | 2008 | Hosp. | 3 |  |  |

- a. Between coverage refers to the average coverage resulting from pairwise comparisons of complete plasmid sequences belonging to different plasmid types (A-F) - b. Within coverage refers to the average coverage resulting from pairwise comparisons of complete plasmid sequences belonging to the same plasmid type.

**Figure 2.**
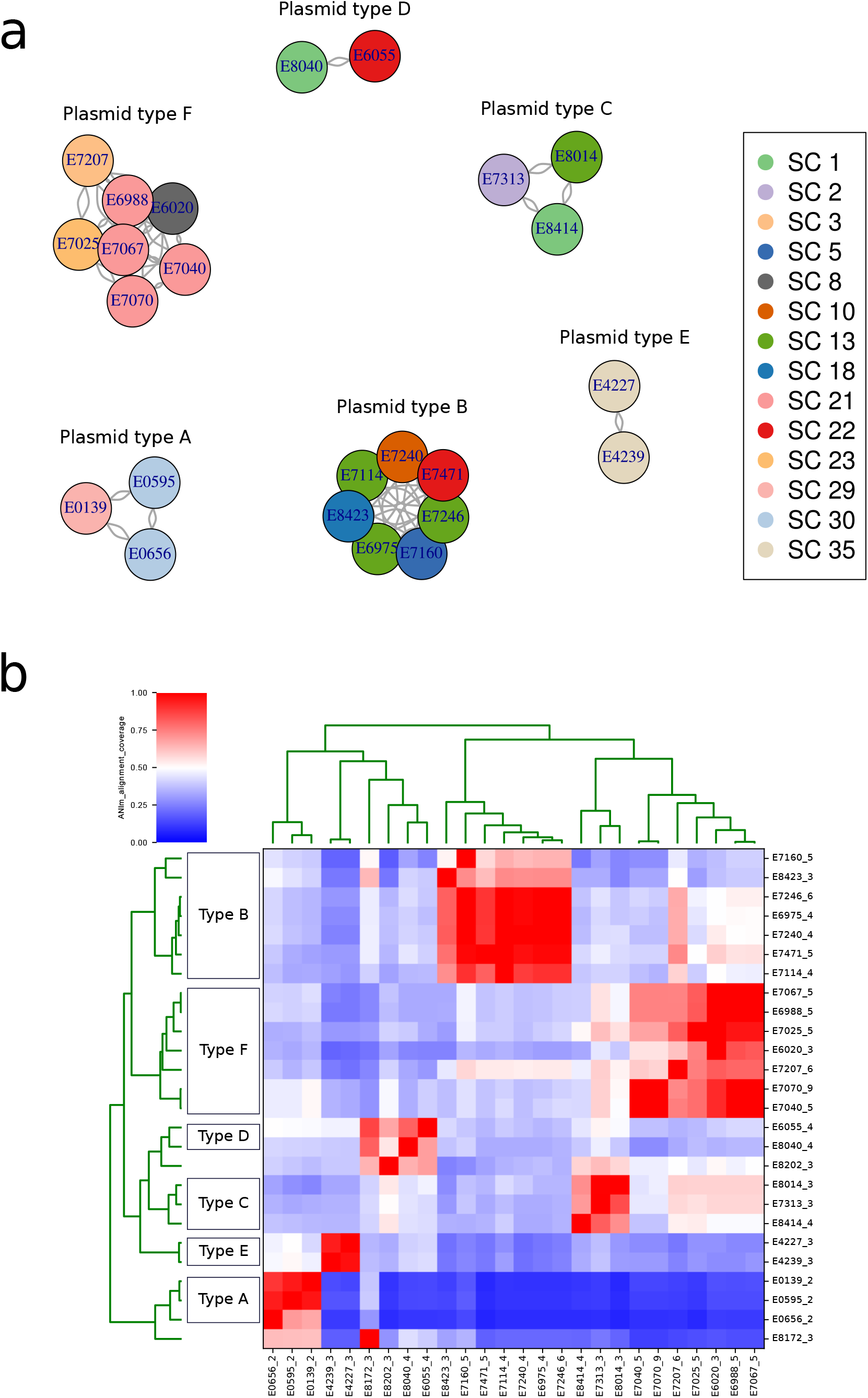
Definition of the plasmid types (A-F) observed in 26 *vanA* complete plasmid sequences. A) Network of *vanA* complete plasmid sequences. Nodes in the graph correspond to plasmids and edges to connections between similar (Mash distance = 0.025, k =21, s =1,000) plasmid sequences. The independent components, present in the network, were designated as plasmid types (n = 6) and nodes were coloured according to the SC isolate carrying the plasmid sequence. B) Heatmap and single-linkage clustering of the pyani pairwise alignment coverage obtained from our set of *vanA* complete plasmid sequences (n = 26). The rectangles present on the right side indicate the grouping of the complete plasmid sequences into the defined plasmid types present in panel A (A-F).

To better understand the modularity and similarity of these plasmid types, we estimated average nucleotide identity (ANI) values using pyani (33). This allowed retrieving ANI coverage and identity values of the aligned regions between two complete plasmid sequences (pairwise comparisons). We observed that the average coverage between plasmid alignments belonging to the same plasmid type was 84% compared to a coverage of 35% when comparing alignments from different plasmid types. The single-linkage hierarchical clustering of the alignment coverage reported by pyani suggested the same plasmid types as inferred in our network approach (Fig. 2B). We did not observe differences in the average identity values between aligned regions within(98.5%) and between (99.7%) plasmid types indicating a common origin of the plasmid modules present in the different types (Fig. S2). To exemplify this, we show the plasmid modularity observed in the complete sequences from plasmid type B, since it had the highest SC diversity and number of associated sequences (Fig. S3).

In Additional File S2, we provide a detailed genomic characterization of the plasmid types (A-F) with a focus on i) replication initiator proteins (rep), ii) Tn*1546* variant compared to the original sequence described by Arthur et al. (30), iii) antimicrobial resistance (AMR) genes distinct from the *vanA* gene cluster and iv) presence of well-known *E. faecium* plasmid TA systems such as ω-ε-ζ and axe-txe (43).

### Plasmid prediction and network integration

Next, we performed a *de-novo* plasmid prediction of the sequences carrying the *vanA* gene cluster in the set of 309 VRE samples using gplas (28). This tool uses a combination of machine-learning and a graph-based approach to predict plasmid sequences from short-read graphs (28). Contigs predicted as belonging to the same plasmid sequence are returned in the same bin.

In 282 isolates (91.2%), the contig which encodes for the *vanA* gene cluster (*vanA* contig) was present in a plasmid bin predicted by gplas. In the remaining isolates (n = 27, 8.7%), the *vanA* contig remained unbinned and thus we could not predict whether the *vanA* gene cluster was part of a plasmid. This could be caused by a fragmented assembly graph due to, for example, a low sequence coverage. Also high differences in the k-mer coverage of contigs from the same plasmid can prohibit binning of plasmid contigs with gplas. The inability of gplas to predict the plasmid location of *vanA* could also be indicative of a chromosomal location of the *vanA* gene cluster. However, manual inspection of the assembly graph in these 27 isolates revealed that the *vanA* k-mer coverage was clearly higher and distinct from the median k-mer coverage of all contigs, indicative of a plasmid location with a higher copy number compared to the chromosome.

Based on these findings, we concluded that in all 309 *vanA* VRE isolates (100%), the gene cluster was present in a plasmid background. The preferential presence of the *vanA* cluster in a plasmid was previously reported by Freitas et al. (53 isolates, 100% plasmid encoded *vanA*)(40) and Wardal et. al (88 isolates, 98% plasmid encoded *vanA*) (41). These 27 isolates in which the *vanA* gene cluster could not be assigned to a particular plasmid bin were excluded from further analysis.

To elucidate gene content and synteny of the plasmid bins (n = 282), we integrated the complete plasmid sequences (Table 1, Figure 2) used to define the six plasmid types A to F with the predicted plasmid bins carrying the *vanA* gene cluster (n = 282). The presence of edges connecting complete plasmids and predicted *vanA* plasmid bins revealed that the predictions had a similar k-mer content and thus further validated the predicted *vanA* plasmid bins (Fig. 3). Furthermore, the distribution of k-mer distances between the plasmid bins (Fig. S5) followed the same pattern as observed with the complete plasmid sequences (Fig. S1). Based on this, the same threshold (0.025) was considered to draw an edge between nodes in the network shown in Figure 3. Nodes were coloured based on the SC of the isolate bearing the complete plasmid sequence or *vanA* plasmid bin (Fig. 3).

**Figure 3.**
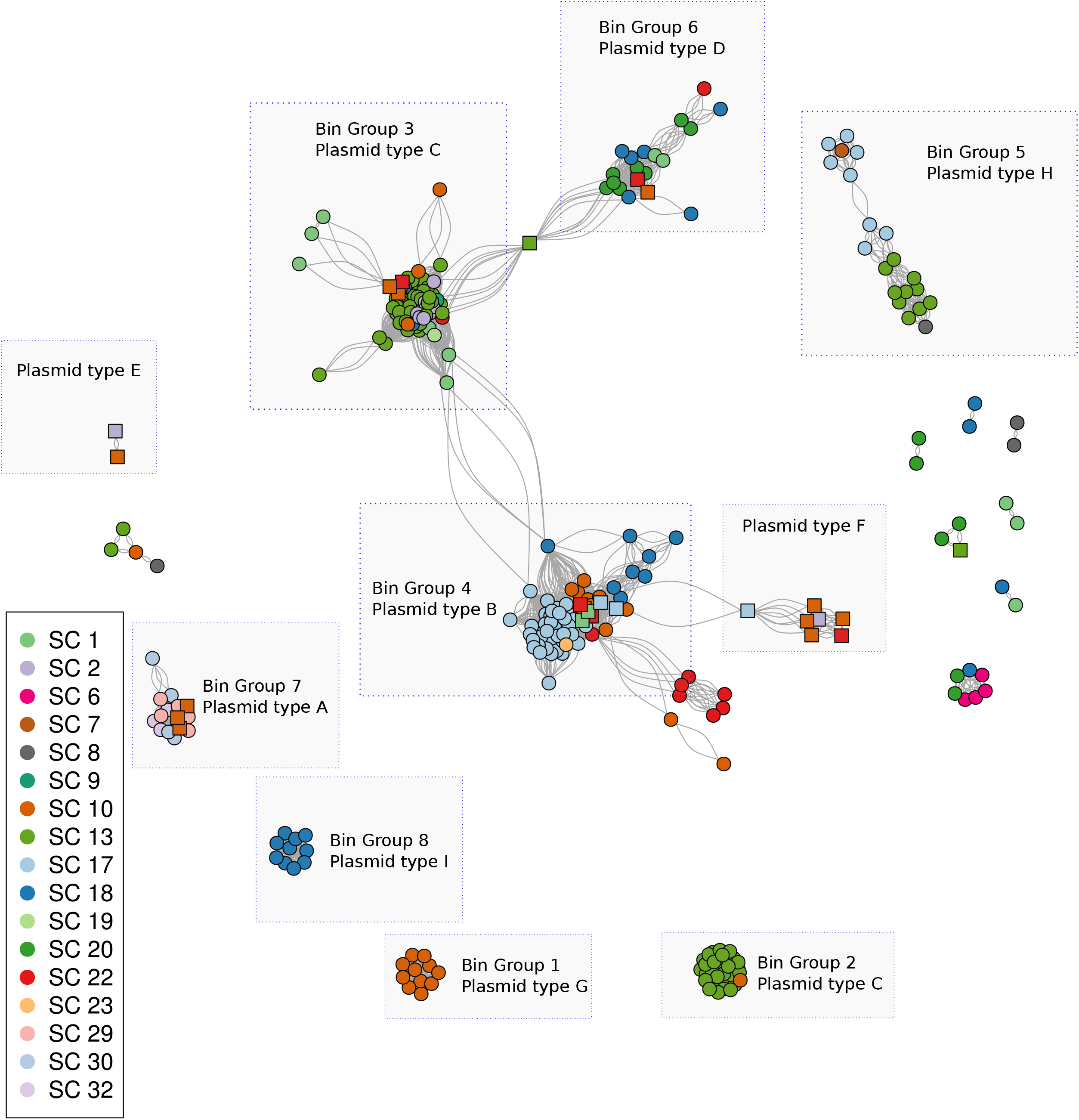
Network of predicted plasmid bins and complete *vanA* plasmid sequences. Nodes corresponding to complete plasmid sequences are highlighted with squared shapes while nodes from predicted plasmid bins are represented with circles. Edges are connections between nodes with similar *vanA* sequences (Mash distance = 0.025, k = 21, s = 1,000). Rectangles indicate the grouping of nodes, based on highly interconnected components, with a size larger than 10 nodes.

We focused on the presence of eight distinct groups of bins which corresponded to graph components that were highly interconnected in the network and had more than 10 nodes (Figure 3, Table 2). Four of these groups co-clustered together with complete plasmid sequences belonging to the plasmid types A, B, C, D. Three groups of plasmid bins (n = 3) were considered as carrying novel plasmid types (G, H, I) since they were not connected to any complete plasmid sequence. This approach allowed assigning 239 isolates with a particular plasmid type (A, B, C, D, G, H, I) (Table 2). In Additional File S2, we provide an extensive characterization of the groups observed in the network (Fig. 3) based on group diversity regarding SC, Tn*1546* variants, geographical site and year of isolation. The Tn*1546* variants found in the different samples with respect to the transposon described by Arthur et. al (30) are outlined in Additional File S3.

**Table 2.**
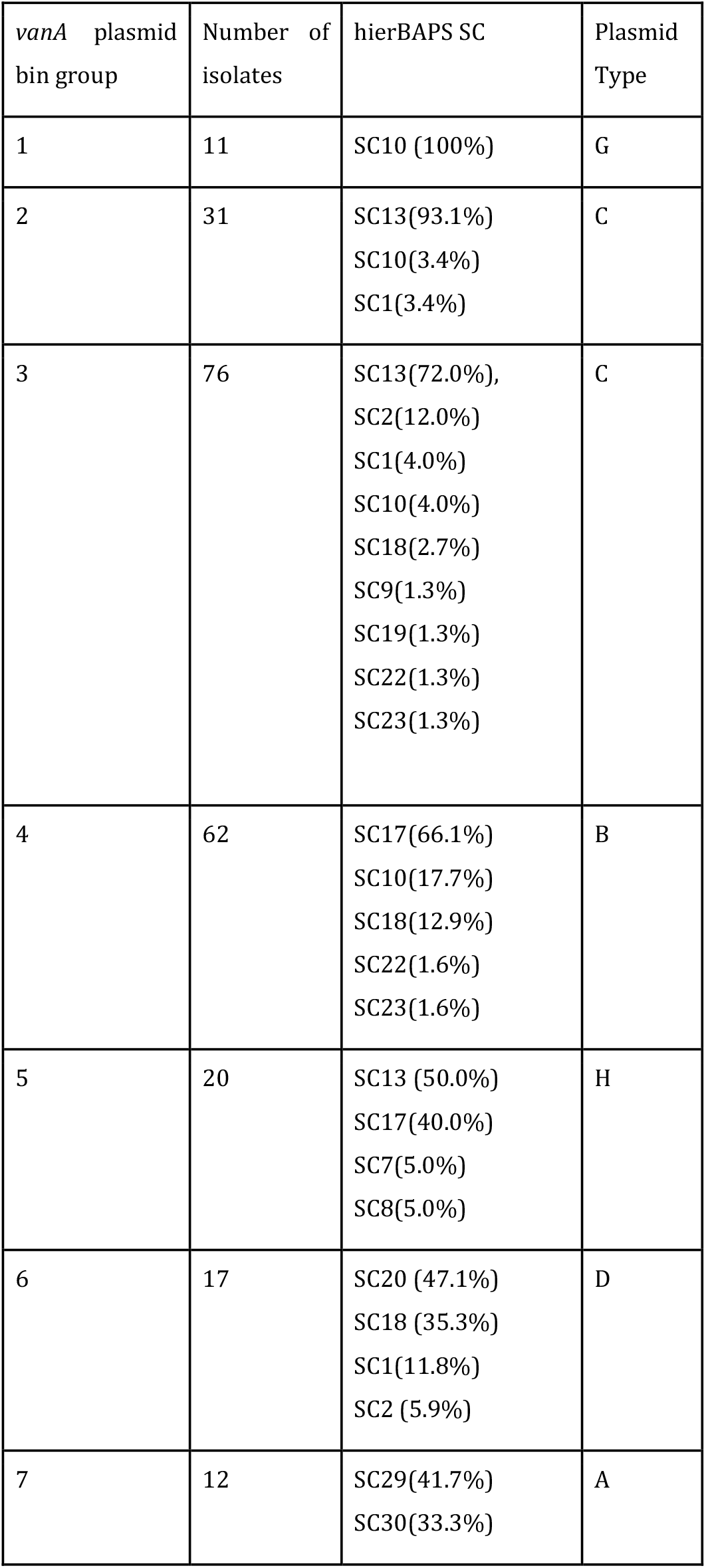
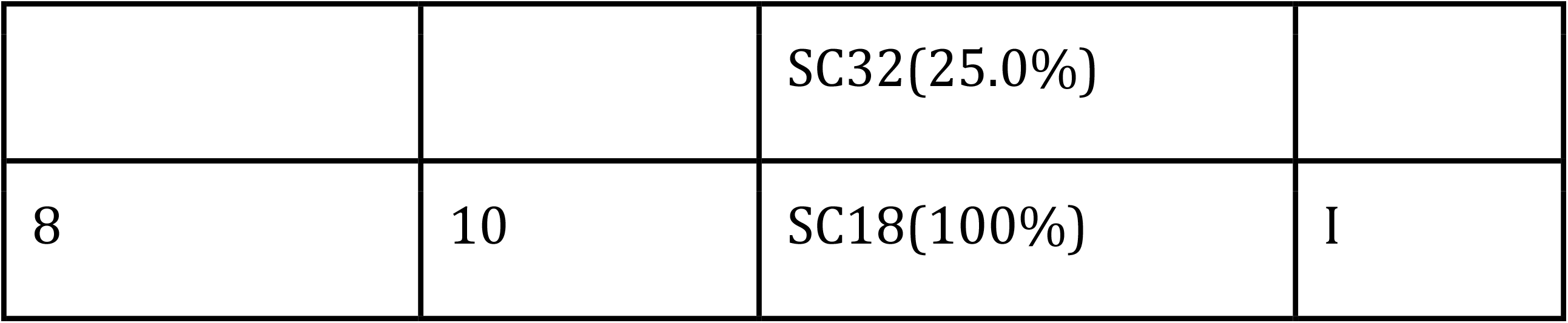
Description of the *vanA* plasmid bin groups (> 10 isolates). For each predicted plasmid type, we indicated the number of isolates forming the predicted plasmid type, percentage of SCs, and assigned plasmid type.

In Figure 4, we combined the core-genome based neighbour-joining tree with SC and *vanA* plasmid type assignments. Some closely related isolates in the core-genome tree had distinct predicted *vanA* plasmid types as exemplified by SC17 containing plasmid types B and H (Fig. 4). The observation of divergence in the plasmid content within the same SC indicates that particular VRE strains horizontally acquired different *vanA* plasmids (Fig. 4). Based on this, we concluded that the plasmid types B, C, D and H were horizontally disseminated between non-clonal *E. faecium* strains whereas the plasmid types A, G and I were linked to isolates which were clonally related (Figure 4, Additional File S2). Plasmid type assignments were also integrated into the Microreact project https://microreact.org/project/FCUD_d1zt to facilitate the exploration of the results.

**Figure 4.**
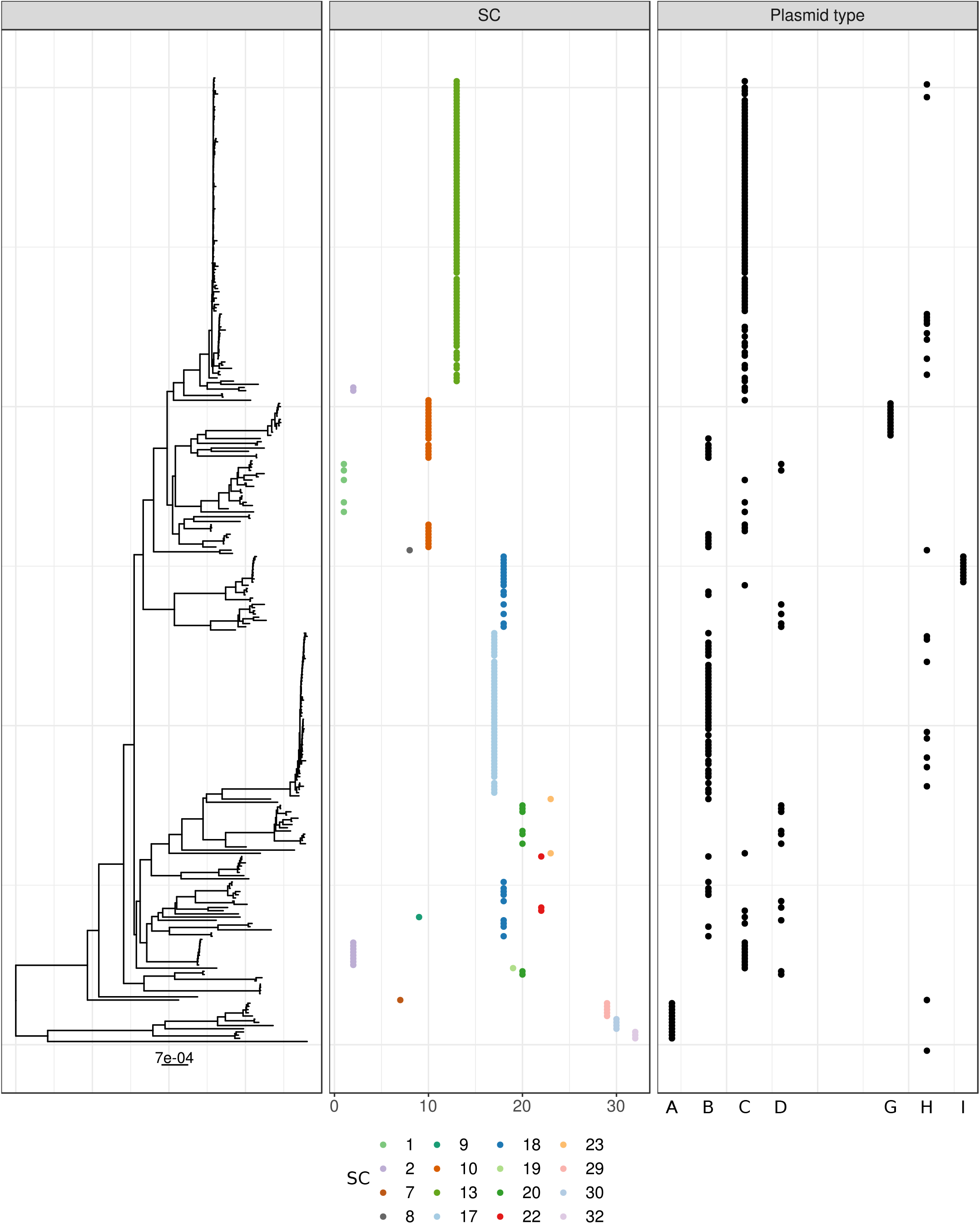
PopPUNK neighbour-joining tree, based on the core genome, in combination with hierBAPS Sequencing Cluster (SC) and *vanA* plasmid type assignments.

### Dynamics of *vanA*-type resistance dissemination

The identification of the seven *vanA* plasmid types (A, B, C, D, G, H, I) present in our Dutch VRE collection allowed us to estimate the contribution of the nested genetic elements, clone (defined as hierBAPS-based SC), plasmid-type and Tn*1546* variant, into the dissemination of the *vanA* gene cluster.

To establish this, we first grouped VRE isolates with a potential epidemiological link which corresponded to isolates from the same Dutch region and recovered within a period of 12 months. To estimate the importance of the inter-regional spread of vancomycin-resistance, the same approach was taken without taking into account the origin of the isolates. We then performed a pairwise “all vs. all” comparison of hierBAPS SC, predicted *vanA* plasmid type and/or Tn*1546* variants of VRE isolates. Based on this, we defined the most likely scenarios of the dissemination of the *vanA* gene cluster: i) clonal dissemination, defined by observing the same SC, *vanA* plasmid type and Tn*1546* variant; ii) plasmid dissemination, defined by observing the same *vanA* plasmid type and Tn*1546* variant but different SC; and iii) Tn*1546* transposition, defined by observing the same Tn*1546* variant in different plasmid types.

We observed that, on average, clonal dissemination was the predominant mode of vancomycin-resistance spread (∼45%), followed by Tn*1546* mobilisation, (∼12%) and plasmid dissemination (∼6%). However, the dynamics of vancomycin-resistance spread were clearly distinct between regions (Fig. 5).

**Figure 5.**
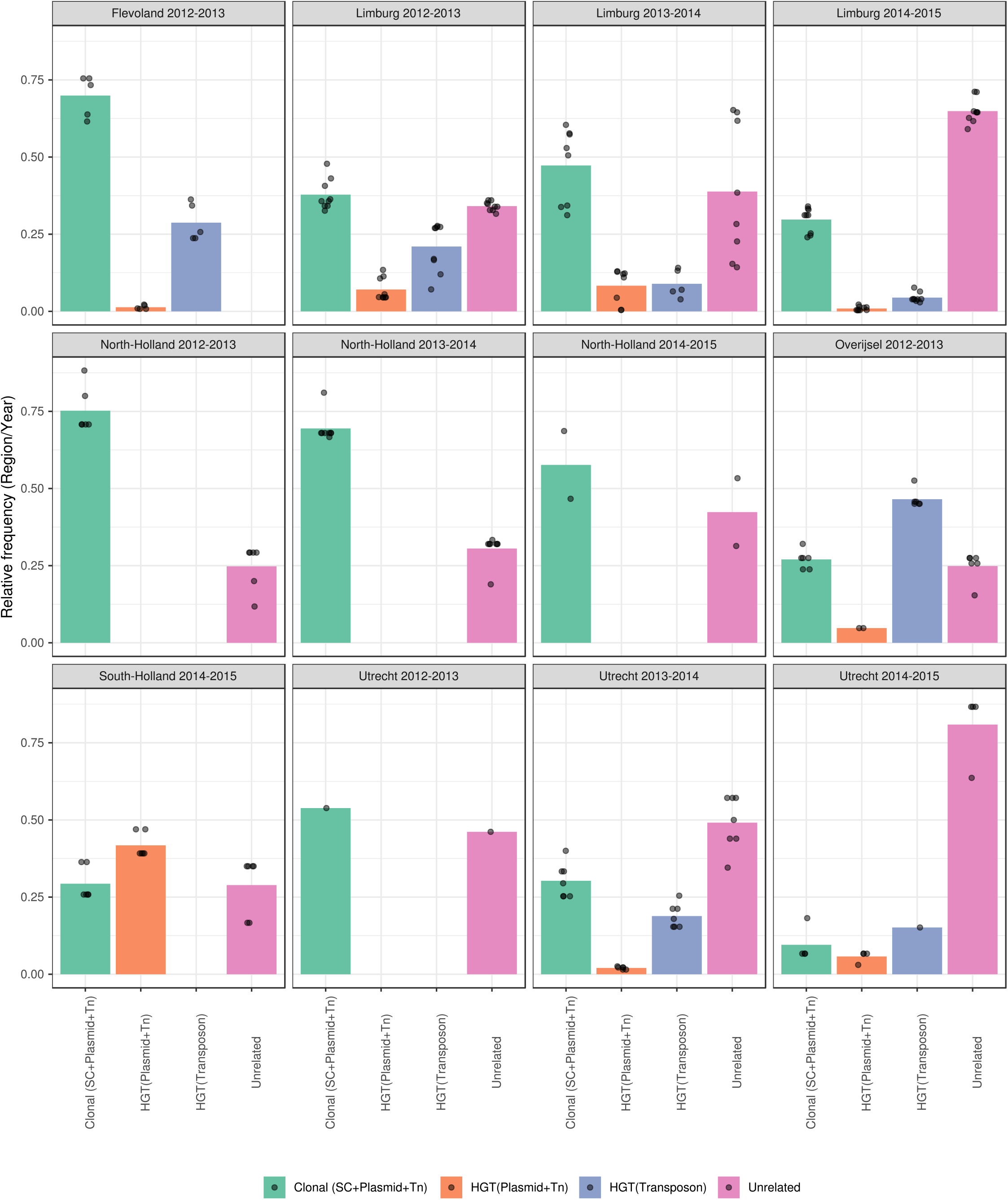
Contribution of nested genomic elements in regional dissemination of *vanA*-type vancomycin resistance in the Netherlands. VRE samples isolated from 12 consecutive months and from the same Dutch region were considered. Clonal dissemination (green bar) corresponded to pairs of isolates sharing the same hierBAPS SC, *vanA* plasmid type and Tn*1546* variant. Plasmid dissemination (orange bar) corresponded to pairs of isolates sharing the same *vanA* plasmid type and Tn*1546* variant but different hierBAPS SC. Dissemination mediated by Tn*1546* transposition (blue bar) corresponded to pairs of isolates sharing the same Tn*1546* variant, different vanA plasmid type and same or different the hierBAPS SC. Unrelated (purple bar) isolates corresponded to pairs of isolates with a different Tn*1546* variant.

Clonal dissemination driven by the hierBAPS SC 17, plasmid type B and Tn*1546* variant represented by the T7658C, G8234T SNPs and deletions in orf1, orf2 (1-3343), contributed most to *vanA*-type vancomycin resistance spread in the provinces of Flevoland (2012-2013, frequency avg. = 70%) and North-Holland (2012-2013, frequency avg. = 75%) (Fig. 5).Interestingly, in North-Holland (2013-2014), clonal dissemination was still the predominant mode of spread (frequency = 70%) but was driven by a different clone defined by the hierBAPS SC 13, plasmid type C, and the Tn*1546* variant with deletions in orf1, orf2 (1-3417) and deletions in the intergenic region 8650-8827. In contrast, mobilisation of the Tn*1546* variant characterised by SNP positions T7658C, G8234T, and deletions in orf1, orf2 (1-3343) between plasmid types B, C, H was predominant in Overijssel (2012-2013, freq = 47%). The Tn*1546* variants present in the different plasmid types are extensively described in Additional File S2.

Lateral transfer of plasmids contributed most to the dissemination of vancomycin resistance in the province of South Holland (2014-2015, freq. avg = 42%). There, we found that plasmid type C together with the Tn*1546* variant with the orf1, orf2 deletions 1-3417, and intergenic region deletions 8650-8827, was found in four distinct clonal backgrounds (SCs: 2, 10, 13, 19). Based on this analysis and the fact that the isolates mainly occurred in the a single hospital of the South Holland region, we conclude that this epidemic rise of vancomycin resistance involved the dissemination of a single plasmid (type C), so could be characterized as a plasmid-driven outbreak of vancomycin-resistance (Fig. S6). We also observed complex scenarios of mixtures of genomic units exemplified by Limburg in 2012-2013, in which clonal (freq = 38%), plasmid dissemination (freq = 7%) and Tn*1546* transposition between distinct plasmid types (freq = 21%), all contributed to the spread of vancomycin-resistance.

Finally, we analysed the spread of vancomycin resistance on a country-wide perspective. Pairwise “all vs. all” comparisons revealed that in most cases VRE strains, plasmid and Tn*1546* variants were unrelated (∼59%). Clonal dissemination was detected in ∼27% of the comparisons. Plasmid dissemination and transposition of Tn*1546* accounted for ∼7% of the comparisons (Fig. S7). However, during the period of 2014-2015, plasmid dissemination increased up to ∼29% which could be linked to the South-Holland plasmid outbreak (2014-2015) described above (Fig. S6).

## Discussion

In this study, we propose a combination of machine-learning and graph-based techniques coupled with a network analysis of the shared plasmid k-mer content to elucidate the dynamics of vertical and horizontal dissemination of AMR genes with short-read WGS. We used this approach to elucidate the mode of *vanA*-type vancomycin resistance dissemination. Clonal and horizontal mobilisation of *vanA* were previously documented but the quantity of these events on a large scale is largely unexplored. Our analysis permitted us to distinguish and quantify dissemination occurring by i) clonal spread, ii) plasmid dissemination and iii) Tn*1546* transposition between distinct plasmid types. This revealed that clonal dissemination was the predominant (∼45%) mode of spread of *vanA*-type of vancomycin resistance, followed by mobilisation due to Tn*1546* transposition among different plasmid types (∼12%) and plasmid dissemination (∼6%). The approach presented here can be applied to study clonal and HGT dissemination of other AMR genes, such as carbapanamese genes (bla^kpc^) in *Enterobacteriaceae* or colistin-resistance genes (*mcr*) in *Escherichia coli*.

Previous studies have described the importance of both clonal and horizontal Tn*1546* dissemination in the emergence of VRE isolates (18,20). However, a quantitative assessment of the contribution from the different nested genomic elements in the dissemination of vancomycin resistance has not been previously performed. Combining existing short-read WGS with complete *vanA* plasmids allowed us to define and characterize several plasmid types present in the collection. The integration of previously completed *vanA* plasmids was essential to elucidate the genetic content of the *vanA* plasmid bins present in our predicted network.These *vanA* plasmid types were defined by ∼99% identity and ∼84% coverage and were present in different clonal backgrounds (SCs), and carried a predominant Tn*1546* variant that accumulated additional SNPs and/or deletions (Additional File S2). The genomic relatedness of strains, using hierBAPS, plasmid types and Tn*1546* variants calling was combined to sketch acomprehensive picture of the molecular epidemiology of *vanA*-type vancomycin resistance in Dutch hospitals.

Transposition of Tn*1546* between different plasmids has been reported before. Heaton et al. showed the transfer of the Tn*1546* element from a non-conjugative to a conjugative plasmid in the same bacterial cell which was mediated by flanking IS*1216* elements (19). Furthermore, horizontal dissemination by larger units than the Tn*1546* as part of a composite transposon has also been previously documented (18,20). The transfer of the Tn*1546* element, alone or as part of a composite transposon could explain the presence of highly similar variants in different plasmid types. Moreover, this observation could also explain the mosaicism observed in the plasmidome of hospitalized patients (44). The inclusion of further complete vancomycin-resistant plasmids into our network could unravel the presence of mosaic plasmids resulting from recombination processes between different vancomycin-resistant plasmid types.

The dissemination of the *vanA* gene cluster can also occur at a plasmid level in which both plasmid type and Tn*1546* variant are horizontally disseminated, as observed in South-Holland between 2014-2015. This type of HGT dissemination can be driven by conjugative plasmids but also from non-conjugative mobilizable plasmids. The presence of non-conjugative plasmids co-residing with conjugative plasmids in the same cell can enhance the horizontal dissemination of both plasmid sequences. This observation has been experimentally validated for the *E. faecium* pHTβ-like plasmid, which contained an efficient conjugation machinery, and allowed the mobilization of other multi-drug resistant and non-conjugative plasmids present in the same cell (45).

The dissemination of vancomycin resistance was previously investigated using WGS in several recent studies (11,46–49) but seldom with a focus on distinguishing clonal and plasmid outbreaks. One exception is the study by Pinholt et al. (14) that used a combination of short-read and long-read sequencing to describe the clonal expansion of VRE in the Capital Region of Denmark between 2012 and 2015. Here, ST80 was defined as responsible for the first observed local outbreaks. These clonal isolates subsequently spread to other hospitals in the same region. The plasmid bearing the *vanA* gene cluster was disseminated to other, non-clonally related vancomycin-susceptible isolates. In our data set covering the same time frame, ST80 represented by SC18 was also a predominant clone (Fig. 1A).

The emergence of *vanA*-type resistance was also investigated in Australia during 2015 using a combination of short and long-read sequencing (15). The study showed the presence of several *vanA* plasmid types which were dominant in each ST group with distinct Tn*1546* variants. This unravelled that, in Australia, the emergence of the *vanA*-type resistance most likely occurred by multiple introductions of different clones which suggested that HGT is not solely responsible for the spread of the *vanA* gene cluster, which is in line with our own results.

Both these studies (14,15), however, followed a reference-based approach to deduce the presence of a particular *vanA* plasmid type. This could mask the presence of plasmid-types which are distinct from the selected reference plasmid(s). In our study, predicted plasmids have been integrated into a network that avoids the arbitrary usage of a reference plasmid and takes plasmid modularity into account. Bipartite networks were previously postulated to explore the pangenome of bacterial species with a particular emphasis on the accessory genome (50). A network approach also allows classifying plasmids in the absence of a common evolutionary history as it can integrate both horizontal and vertical inheritance, in contrast to phylogenetic trees (51,52). The classification of plasmids based on k-mer similarity networks has also recently been proposed by Acman et al. (53). Consequently, our network-based analysis could be expanded to include other *vanA* plasmids from plasmid databases such as Plasmid Atlas or PLSDB (54,55) and could provide a global picture of the dissemination of vancomycin-resistance.

A focus on the core genome can overestimate the number of isolates that are considered as non-related and thus missing potential epidemiological links. In line with Harris et al. (56), we encourage the shift from a traditional core genome view on outbreak investigations to a new perspective that also includes the analyses of HGT mobilisation of AMR genes to effectively confirm potential epidemiological links and correctly evaluate the effectiveness of infection control policies. We showed that highly similar plasmids can be transferred between different SC’s which challenges the interpretation of AMR outbreak studies that are solely focused on core-genome analysis. A factor that contributes to this clonality centricity are the limitations inherent to short-read WGS from which the assembly of plasmids is difficult and error-prone due to the high number of repeated sequences (57). For full resolution, many studies recommend long-read sequencing to complete chromosomes and plasmids (16,57,58). We show in this study, however, that distinguishing between different modes of spread is feasible also in the absence of long-read data.

A limitation of this work is that only a single colony per isolate was sequenced. This masked the true underlying diversity present in the bacterial population and could have prohibited the detection of potential epidemiological links. While our data set allowed us to define the most likely events of dissemination, we lacked patient admission and ward movement information to discern transmission routes and events, as previously shown by Raven et al. (11) and Neumann et al. (59). The absence of data on vancomycin-susceptible isolates also prevented us to deduce how the distinct plasmid types and Tn*1546* variants were introduced into the Dutch hospitals. Nonetheless, we succeeded to provide one of the first quantitative assessments to discern the dynamics and contribution of clonal and horizontal transmission in the dissemination of vancomycin resistance.

## Conclusions

This study has shown that clonal dissemination was the preferential mode of dissemination of *vanA*-type vancomycin resistance in Dutch hospitals between 2012 and 2015. However, we also detected outbreak settings in which HGT dissemination, either mediated by plasmid or Tn*1546* dissemination, contributed most to the spread of resistance. Our analyses showed the importance of taking all nested genomic elements into account to effectively elucidate how resistance spreads in healthcare settings. This is fundamental to corroborate potential epidemiological links that could be neglected by uniquely considering strain relatedness. Only then, the effectiveness of current infection control policies to prevent AMR spread can be truly assessed.

## Data Availability

The raw paired-end reads of the 309 VRE isolates are available through the European Nucleotide
Archive project PRJEB28495. The complete *vanA* plasmid sequences (n = 26) used to define the
plasmid types (A-F) are also available through the NCBI Bioproject PRJEB28495 and the gitlab
project https://gitlab.com/sirarredondo/vancomycin_dissemination
The complete code used to generate the results present in this manuscript is provided in a
RMarkdown document available through
https://gitlab.com/sirarredondo/vancomycin_dissemination.

## List of abbreviations

WHO: World Health Organization
VRE: Vancomycin resistant *Enterococcus faecium*
WGS: Whole-genome sequencing
HGT: Horizontal gene transfer
AMR: Antimicrobial resistance
SC: Sequencing cluster
SNP: Single nucleotide polymorphism
ORF: Open reading frame

## Declarations

### Ethics approval and consent to participate

Not applicable.

### Consent for publication

Not applicable.

### Availability of data and materials

The raw paired-end reads of the 309 VRE isolates are available through the European Nucleotide Archive project PRJEB28495. The complete *vanA* plasmid sequences (n = 26) used to define the plasmid types (A-F) are also available through the NCBI Bioproject PRJEB28495 and the gitlab project https://gitlab.com/sirarredondo/vancomycin_dissemination

The complete code used to generate the results present in this manuscript is provided in a RMarkdown document available through https://gitlab.com/sirarredondo/vancomycin_dissemination.

### Authors’ contributions

AS and RW initiated and supervised the project. SA performed the computational analysis. SA, AS and RW wrote the manuscript. All authors read and commented on the manuscript. All authors read and approved the final manuscript.

## Acknowledgments

Not applicable.

## Additional files

Additional File S1 (.csv) - Metadata table related to the 309 VRE isolates.

Additional File S2 (.pdf) - Supplementary Results. Characterization of the genomic organization present in the plasmid types. Characterization of the predicted plasmid bins.

Additional file S3 (.csv) - TETyper output table with the description of the Tn*1546* variants found in all VRE isolates respect to the original Tn*1546* element (NCBI Nucleotide Accession M97297).

Additional File S4 (.pdf) - Supplementary Figures (Fig S1-Fig S7).

